# Comparing in-person, blended and virtual training interventions; a real-world evaluation of HIV capacity building programs in 16 countries in sub-Saharan Africa

**DOI:** 10.1101/2023.02.08.23285641

**Authors:** E Kiguli-Malwadde, M Forster, A Eliaz, J Celentano, E Chilembe, ID Couper, ET Dassah, MR De Villiers, O Gachuno, C Haruzivishe, J Khanyola, S Martin, K Motlhatlhedi, R Mubuuke, K A Mteta, P Moabi, A Rodrigues, D Sears, F Semitala, D von Zinkernagel, MJA Reid, F Suleman

## Abstract

**Introduction:** We sought to evaluate the impact of transitioning a multi-country HIV training program from in-person to online by comparing digital training approaches implemented during the pandemic with in-person approaches employed before COVID-19.

**Methods:** We evaluated mean changes in pre-and post-course knowledge scores and self-reported confidence scores for learners who participated in (1) in-person workshops (between October 2019 and March 2020), (2) an entirely asynchronous, Virtual Workshops [VW] (between May 2021 and January 2022), and (3) a blended Online Course [OC] (between May 2021 and January 2022) across 16 SSA countries. Learning objectives and evaluation tools were the same for all three groups.

**Results:** Across 16 SSA countries, 3023 participants enrolled in the in-person course, 2193 learners participated in the virtual workshop and 527 in the online course. The proportions of women who participated in the VW and OC were greater than the proportion who participated in the in-person course (60.1% and 63.6%, p<0.001). Nursing and midwives constituted the largest learner group overall (1145 [37.9%] vs. 949 [43.3%] vs. 107 [20.5%]).

Across all domains of HIV knowledge and self-perceived confidence, there was a mean increase between pre- and post-course assessments, regardless of how training was delivered. The greatest percent increase in knowledge scores was among those participating in the in-person course compared to VW or OC formats (13.6% increase vs. 6.0% and 7.6%, p<0.001). Gains in self-reported confidence were greater among learners who participated in the in-person course compared to VW or OC formats, regardless of training level (p<0.001) or professional cadre (p<0.001).

**Conclusions:** In this multi-country capacity HIV training program, in-person, online synchronous and blended synchronous/asynchronous strategies were effective means of training learners from diverse clinical settings. Online learning approaches facilitated participation from more women and more diverse cadres. However, gains in knowledge and clinical confidence were greater among those participating in in-person learning programs.

## INTRODUCTION

Even before the COVID-19 pandemic, there was a pressing need to expand the health workforce and optimize team-based care across sub-Sharan Africa (SSA), especially in settings of scarce health resources.(1) While causing unprecedented disruptions to clinical care and health professions training opportunities, the COVID-19 pandemic presented a unique opportunity to rapidly scale new training modalities, including maximizing the use of online learning platforms.(2, 3) Numerous African and international training programs developed and implemented remote educational programs to address safety concerns regarding COVID-19 transmission and to ensure continuity of training programs in spite of government interventions to reduce infection and transmission.(4, 5) One such example was the Strengthening Inter-professional Education in HIV (STRIPE HIV) initiative, which rapidly transitioned its in-person training program to online.(1) STRIPE HIV is an HIV training program funded by the United States government through the President’s Emergency Plan for AIDS Relief (PEPFAR), that includes a diverse network of health professional training institutions across SSA, led by the African Forum for Research and Education in Health (AFREhealth) and the University of California, San Francisco.(6) At the start of the COVID-19 pandemic, the STRIPE team rapidly implemented remote educational programming, including developing a learning management system (LMS) accessible to health professions at affiliated training institutions across SSA.(2) The internet-based educational programs consisted of inter-professional HIV training content for pre-service and in-service learners from diverse professional cadres; while the format was altered to accommodate online learning, the curriculum was the same as that employed pre-COVID.(1, 6)

In advance of implementing this online approach to training, limited research comparing the effectiveness of in-person and remote educational formats across SSA had been undertaken.(7) Previous research has evaluated the impact of numerous capacity building interventions to target individual healthcare provider behavior in low and middle-income countries (LMICs),(7, 8, 9, 10) but very little research has evaluated the comparative benefits of online capacity building interventions in such settings. The in-person and online remote educational programs developed by STRIPE HIV and AFREhealth provide a unique opportunity to evaluate whether transitioning to online learning significantly impacted inter-professional training in HIV care. As such, we sought to compare the impact of the in-person and online formats of inter-professional education on knowledge and confidence among learners of diverse cadres across fourteen countries in SSA.

## METHODS

### Curriculum design and implementation: Year One In-Person Program

A case-based curriculum in inter-professional HIV training was developed and implemented during year one (Y1) of STRIPE HIV’s educational program, previously described.(1) The program was developed using Kern’s six-step approach by a collaborative team of local and international HIV practitioners and health professions education experts.(11) The inter-professional HIV training included 17 case-based modules with at least four required modules delivered in person over a two-day workshop. The program was initially implemented across 14 countries in SSA between October 2019 and April 2020.

### Curriculum design and implementation: Year Two Online Synchronous Program

In year two (Y2), which coincided with the spread of COVID-19 across SSA, participating academic institutions opted for one of two modalities, based on Internet access and existing expertise using educational technologies. Many schools opted to pursue training using a Virtual Workshop (VW) format, which consisted of synchronous online workshops. This approach utilized a revised version of the in-person training content from Y1, adapted for delivery over Zoom™ and taught by both local and international facilitators, but incorporating both small-group activities and large-group discussions like the in-person approach. Learners from fourteen countries, including learners from several countries that had not previously (Algeria, Madagascar, Mozambique, and Somalia), participated in the Virtual Workshop beginning in May 2021.

### Curriculum design and implementation: Year Two Online Blended Program

Some institutions opted to deliver the same modular content using an online approach that included blended asynchronous and synchronous content (hitherto referred to as the Online Course [OC]). The asynchronous content in the blended program was completed independently online using a Moodle-based learning management system. After completion of the asynchronous content, learners took part in synchronous inter-professional sessions carried out on Zoom™. Learners participated in a synchronous session associated with each of the modules using a flipped classroom approach.(12) The Online Course was implemented across six countries in SSA beginning in June 2021.

### Study Design & Subjects

We compared learner knowledge and confidence across the three educational programs: the In-Person Training (Y1), the Virtual Workshop (Y2), and the Online Course (Y2). Subjects were learners who completed the STRIPE registration survey, as well as pre-test and post-test knowledge and confidence assessments. We evaluated self-reported learner demographics including gender identity, training level, health profession, and country of residence. Pre-service learners were defined as learners enrolled in a health profession training institution and working towards their degree. In-service or post-graduate, clinical providers were learners who had already graduated from their health professions training program and engaged in clinical practice. Additionally, we defined early in-service learners as learners who had graduated from training fewer than 12 months prior to participating in the STRIPE HIV course.

To compare the three learning approaches, we evaluated data from four training four modules that utilized the same pre-test and post-test assessments across the three educational formats. The four case-based modules were titled *New HIV Diagnosis and Antiretroviral Therapy (ART) Initiation in a Woman of Childbearing Age, Management of HIV-TB Co-Infection, PMTCT & Care for the Pregnant Woman with HIV*, and *Care for the Paediatric Patient with HIV*. The specific knowledge and confidence assessment questions from each module utilized in the analysis are shown in Supplemental Table 1.

### Knowledge Assessment

We evaluated learning among participants by assessing the mean difference in pre-test and post-test knowledge scores and percent change in knowledge scores. For Y1, data were collected during the period of training from October 2019 to March 2020. For Y2, data were collected from May 2021 to January 2022. We compared mean knowledge differences and percent increase across the educational programs and stratified knowledge differences by health profession and training level.

### Confidence Assessment

We evaluated confidence among participants by assessing the mean difference in pre-test and post-test confidence scores in three arenas: clinical confidence, confidence in working as part of an inter-professional team (IP Confidence), and confidence in implementing quality improvement strategies (QI Confidence). We then compared mean differences in confidence scores across the three educational programs, stratified by health profession and training level.

### Statistical Analysis

We compared learner demographics using Pearson chi-squared tests. We utilized Analysis of Variance (ANOVA) and Tukey’s Honest Significance tests to compare mean differences in knowledge and confidence across the three educational programs. Analyses were carried out using RStudio (2022.07.1+554). Additionally, *p*<0.01 was considered statistically significant.

### Ethics Statement

The design of the training program, including the topics covered and the format of the training, was informed by input from focus-group discussions with patient groups, learners (both pre-service and early career professionals) and HIV educators from a variety of settings in SSA, and has been previously described.(6, 13) Assessment tools, evaluating learners knowledge and confidence, were also piloted with a subset of multidisciplinary learners before the full program was launched. All learners were given access to their pre and post score test results, via the program’s website. In addition, aggregate, site-level evaluation data were also posted on the program’s website. The protocol for this project was reviewed and approved by the University of California, San Francisco’s Institutional Review Board (IRB) in San Francisco, California. Verbal consent was required at the time of participation in the study as approved by the IRB (protocol #: 19–28,447).

## FINDINGS

### Learner Demographics

During Y1, from October 2019 to March 2020, 5027 participants from 12 countries in SSA participated in the In-Person Course, and 3023 of those learners completed pre-test and post-test assessments. During Y2, from June 2021 to November 2022, 7107 learners from 15 countries in sub-Saharan Africa enrolled in the two remote educational programs and 3477 of those learners completed the pre-test and post-test assessments. Among the 3477 remote learners included in the study, 2595 completed the VW training and 628 completed the OC training (Table 1). Complete demographic data was available for 3022, 2193 and 527 learners in the In-person, VW and OC respectively, while pre- and post-course evaluative data was available for 3027, 2595 and 629 participants in each of these groups.

**Table 1.** Demographic summary of Y1 in-person, Y2 Virtual Workshop, and Y2 Online Course learners included in the study

|  | Y1 In-Person |  | Y2 Virtual Workshop |  | Y2 Online Course |  |
| --- | --- | --- | --- | --- | --- | --- |
|  | No. | (%) | No. | (%) | No. | (%) |
| **Gender Identity** | 3023 | (100.0) | 2193 | (100.0) | 527 | (100.0) |
| Female | 1570 | (51.9) | 1319 | (60.1) | 335 | (63.6) |
| Male | 1281 | (42.4) | 873 | (39.8) | 192 | (36.4) |
| Additional | 172 | (5.7) | 1 | (0) | 0 | (0) |
| **Current Training Level** | 3022 | (100.0) | 2193 | (100.0) | 523 | (100.0) |
| Pre-service learner | 1755 | (58.1) | 1518 | (69.2) | 299 | (57.1) |
| Post-graduate new provider* | 727 | (24.1) | 291 | (13.3) | 66 | (12.6) |
| Post-graduate provider** | 540 | (17.9) | 384 | (17.5) | 158 | (30.2) |
| **Current Health Profession** | 3023 | (100.0) | 2193 | (100.0) | 523 | (100.0) |
| Laboratory | 365 | (12.1) | 86 | (3.9) | 117 | (22.4) |
| Medical | 902 | (29.8) | 582 | (26.5) | 140 | (26.8) |
| Nursing/midwifery | 1145 | (37.9) | 949 | (43.3) | 107 | (20.5) |
| Pharmacy | 312 | (10.3) | 373 | (17.0) | 95 | (18.2) |
| Other | 299 | (9.9) | 203 | (9.3) | 64 | (12.2) |
| **Country** | 3022 | (100.0) | 2595 | (100.0) | 628 | (100.0) |
| Algeria | 0 | (0) | 14 | (0.5) | 3 | (0.5) |
| Botswana | 174 | (5.8) | 2 | (0.1) | 113 | (18.0) |
| Ethiopia | 50 | (1.7) | 54 | (2.1) | 0 | (0) |
| Ghana | 733 | (24.3) | 542 | (20.9) | 0 | (0) |
| Kenya | 1 | (0.0) | 326 | (12.6) | 1 | (0.2) |
| Lesotho | 130 | (4.3) | 686 | (26.4) | 0 | (0) |
| Madagascar | 0 | (0) | 1 | (0) | 0 | (0) |
| Malawi | 512 | (16.9) | 254 | (9.8) | 73 | (11.6) |
| Mozambique | 0 | (0) | 17 | (0.7) | 0 | (0) |
| Nigeria | 192 | (6.4) | 0 | (0) | 252 | (40.1) |
| Somalia | 0 | (0) | 1 | (0) | 0 | (0) |
| South Africa | 323 | (10.7) | 128 | (4.9) | 1 | (0.2) |
| Tanzania | 50 | (1.7) | 194 | (7.5) | 0 | (0) |
| Uganda | 635 | (21.0) | 0 | (0) | 0 | (0) |
| Zambia | 110 | (3.6) | 133 | (5.1) | 0 | (0) |
| Zimbabwe | 112 | (3.7) | 243 | (9.4) | 186 | (35.6) |

Y1: year one; Y2: year two. \*New post-graduate provider: provider participating in training less than 12 months since graduating from health profession training program; \*\*post-graduate provider: provider participating in training at least 12 months or more since graduating from health profession training program.

### Gender

Among Y1 participants, 1570 (51.9%) identified as female, 1281 (42.4%) identified as male, and 172 (5.7%) identified as other. The proportions of female learners in the Y2 Virtual Workshop and Y2 Online Course were significantly greater than the Y1 in-person program (60.1% [n=1319] and 63.6% [n=335] vs. 51.9% [n=1570], p<0.001).

### Training Level

Among learners in the Y1, 1755 (58.1%) identified as pre-service, 727 (24.1%) identified as new providers, less than 12 months from graduation, and 540 (17.9%) identified as post-graduate providers, more than 12 months from graduating their training program (Table 1). The proportions of in-service new providers in the Y2 Virtual Workshop and Y2 Online Course were significantly lower than the Y1 in-person program (13.3% [n=291] and 12.6% [n=66] vs 24.1% [n=727], p<0.001).

### Health Professions

In both the Y1 In-person program and remote Y2 educational programs, at least four different health professions participated in the training. Health professions included laboratory, medical, nursing/midwifery, pharmacy, and other, (We included physical therapists and nutritionists in the “other” category given the small number of learners from those professional cadres.) Nursing/midwifery learners constituted the largest proportion of learners in the In-person (n=1145 [37.9%]) and VW (n=949 [43.3%]) courses, while medical providers constituted the largest group in the OC (n=140 [26.8%]).

### Knowledge Assessment

Across all subgroups of learners, HIV knowledge scores increased between the pre- and post-course tests. The percent increase in knowledge scores was significantly lower for those who participated in either the VW or OC formats than among learners who participated in the Y1 In-person training (+6.0% and +7.6% vs +13.6%, p<0.001) (Table 2). Moreover, across the three training levels, the increase in knowledge scores was greatest for participants in the Y1 In-person course compared to either VW or OC. Among pre-service and post-graduate new provider learners there were no significant differences in knowledge scores noted between VW and OC participants, but there were greater gains in knowledge among post graduate provider learners who participated in the OC compared to the VW (Table 2).

**Table 2.** Total learner mean knowledge score differences during Y1 in-person Training, Y2 Virtual Workshop, and Y2 Online Course by Total Score, Training Level, and Health Profession compared using ANOVA

|  | Y1 In-person<br>Mean (%) Difference |  | Y2 Virtual Workshop<br>Mean Difference |  | Y2 Online Course<br>Mean Difference |  | P-Value* |
| --- | --- | --- | --- | --- | --- | --- | --- |
|  | n | mean (%) | n | mean (%) | n | mean (%) |  |
| Total Score | 3027 | +1.2 (13.6) | 2595 | +0.5 (6.0) | 629 | +0.7 (7.6) | <0.001 |
| Training Level |  |  |  |  |  |  |  |
| Pre-service learner | 1755 | +1.3 (14.1) | 1518 | +0.5 (5.1) | 299 | +0.6 (7.0) | <0.001 |
| Post-graduate new provider | 727 | +1.2 (13.4) | 291 | +0.6 (6.5) | 66 | +0.7 (7.4) | <0.001 |
| Post-graduate provider | 540 | +1.1 (12.5) | 384 | +0.7 (7.6) | 158 | +1.0 (11.6) | <0.001 |
| Health Profession |  |  |  |  |  |  |  |
| Laboratory | 365 | +1.7 (19.4) | 86 | +0.6 (6.8) | 117 | +1.2 (13.7) | <0.001 |
| Medical | 902 | +1.0 (10.7) | 582 | +0.5 (5.7) | 140 | +0.8 (9.3) | <0.001 |
| Nursing/midwifery | 1145 | +1.4 (15.0) | 949 | +0.5 (5.8) | 107 | +0.4 (4.5) | <0.001 |
| Pharmacy | 312 | +1.1 (12.3) | 373 | +0.5 (5.0) | 95 | +0.6 (6.5) | <0.001 |
| Other | 299 | +1.0 (11.6) | 203 | +0.5 (5.9) | 64 | +0.6 (6.7) | <0.01 |

\*Analysis of Variance (ANOVA) test used to compare mean differences in knowledge across the three educational programs. Abbreviations: Y1: year one; Y2: year two; post-graduate new provider: provider participating in training less than 12 months since graduating from health profession training program; Post-graduate provider: provider participating in training at least 12 months or more since graduating from health profession training program; % diff = Percent difference between the mean pre and the mean post score. Maximum Score = 9.

Regardless of health professions cadre, gains in knowledge were greater among Y1 In-person learners compared to learners who participated in the VW or OC formats. Among learners from nursing, pharmacy and other cadres, there was no difference in knowledge gains between participants in the VW and OC formats.

### Confidence Assessment

Across all subgroups of learners, self-perceived confidence increased between the pre- and post-course assessments (Table 3, Figures 1-2). Overall, the percent increases in confidence scores were significantly lower among learners who participated in the VW or OC than among learners who participated in the Y1 in-person training (+15.0% and +14.3% vs. +18.9%, p<0.001). Gains in confidence were significantly greater among pre-service learners (p<0.001) and post-graduate new provider learners (p<0.001) who participated in Y1 in-person training compared to the VW or OC programs (Figure 1); differences in confidence between the three programs were not significantly different among post-graduate provider learners (p=0.053). Increases in confidence were greater among medical, nursing/midwifery, and pharmacy learners participating in Y1 in-person training compared to the VW or OC formats (p=0.016, p<0.001 and p<0.001 respectively); differences in confidence among laboratorian participants between the three course were not significantly different (p=0.876) (Figure 2). Regardless of the domain of confidence assessed ((i) clinical confidence, (ii) confidence to engage in Inter-professional practice (IP), and (iii) confidence in quality improvement practice (QI)) there was no significant difference in gains in confidence between participants in the VW or OC formats (Supplemental Figure).

**Table 3.** Total learner mean confidence score differences during Y1 in-person Training, Y2 Virtual Workshop, and Y2 Online Course by Total Score, Training Level, Health Profession, and Confidence Type compared using ANOVA

|  | Y1 In-person<br>Mean (%) Difference |  | Y2 Virtual Workshop<br>Mean Difference |  | Y2 Online Course<br>Mean Difference |  | P-Value* |
| --- | --- | --- | --- | --- | --- | --- | --- |
|  | n | mean (%) | n | mean (%) | n | mean (%) |  |
| Total Score | 3027 | +13.6 (18.9) | 2595 | +10.8 (15.0) | 629 | +10.3 (14.3) | <0.001 |
| Training Level |  |  |  |  |  |  |  |
| Pre-service learner | 1755 | +13.6 (18.9) | 1518 | +9.9 (13.8) | 299 | +11.0 (15.3) | <0.001 |
| Post-graduate new provider | 727 | +14.3 (19.8) | 291 | +11.0 (15.2) | 66 | +10.6 (14.8) | <0.001 |
| Post-graduate provider | 540 | +12.8 (17.8) | 384 | +11.7 (16.3) | 158 | +10.4 (14.4) | 0.053 |
| Health Profession |  |  |  |  |  |  |  |
| Laboratory | 365 | +10.4 (14.5) | 86 | +11.0 (15.3) | 117 | +11.0 (15.2) | 0.876 |
| Medical | 902 | +15.7 (21.8) | 582 | +14.4 (20.0) | 140 | +13.1 (18.2) | 0.016 |
| Nursing/midwifery | 1145 | +14.2 (19.8) | 949 | +8.5 (11.8) | 107 | +7.5 (10.4) | <0.001 |
| Pharmacy | 312 | +13.4 (18.6) | 373 | +9.6 (13.3) | 95 | +12.0 (16.6) | <0.001 |
| Other | 299 | +9.2 (12.8) | 203 | +8.9 (12.3) | 64 | +9.1 (12.8) | 0.969 |
| Confidence Type |  |  |  |  |  |  |  |
| Clinical | 3027 | +11.1 (18.5) | 2595 | +8.8 (14.7) | 629 | +8.3 (13.9) | <0.001 |
| IP | 3027 | +1.4 (16.9) | 2595 | +1.1 (14.3) | 629 | +1.2 (14.6) | <0.001 |
| QI | 3027 | +1.1 (28.7) | 2595 | +0.8 (21.2) | 629 | +0.8 (19.5) | <0.001 |

**Figure 1.**
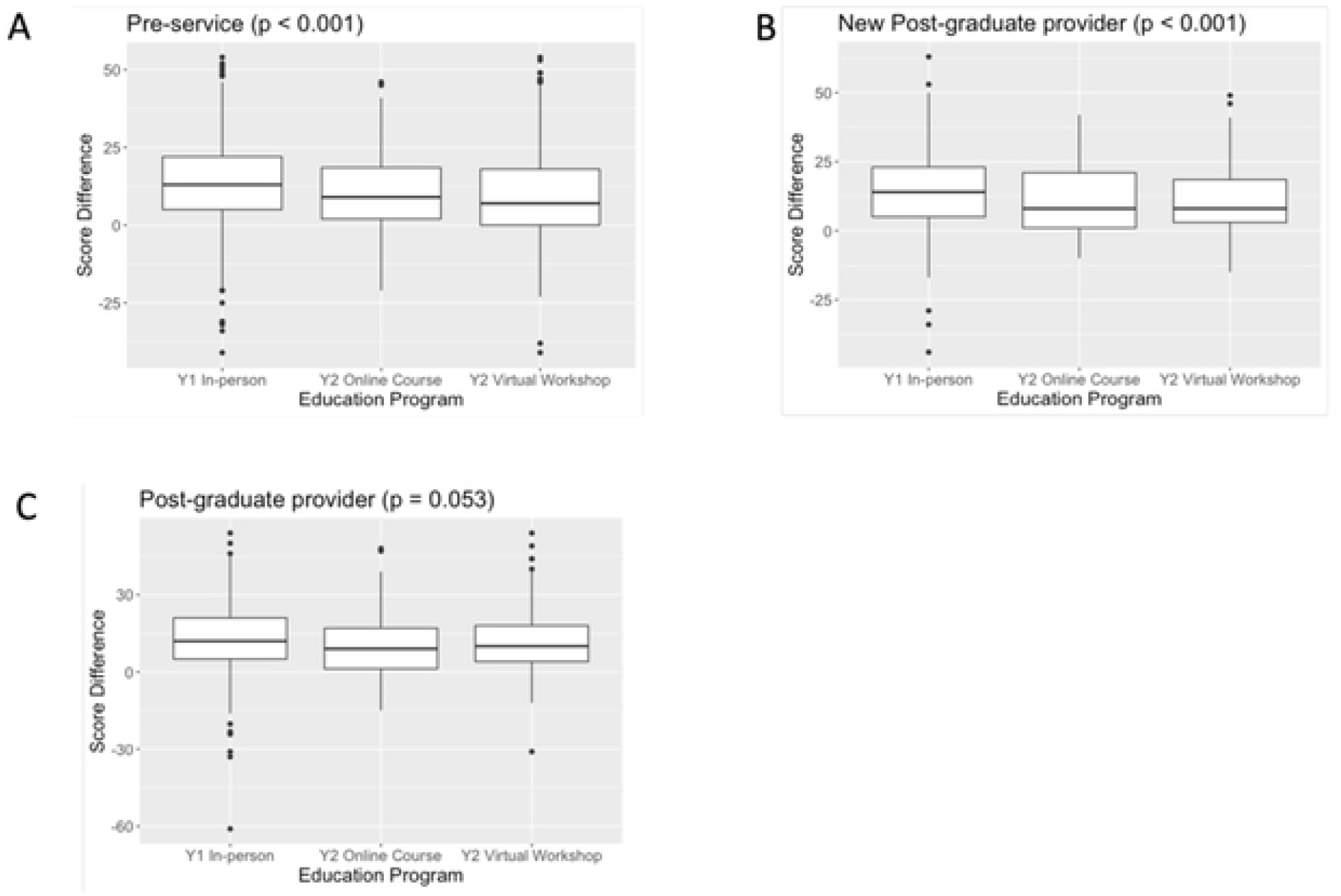
Total confidence score difference by education program for Pre-service Learner (A, n = 1755), New Post-graduate provider (B, n = 727), and Post-graduate provider (C, n = 540).

**Figure 2.**
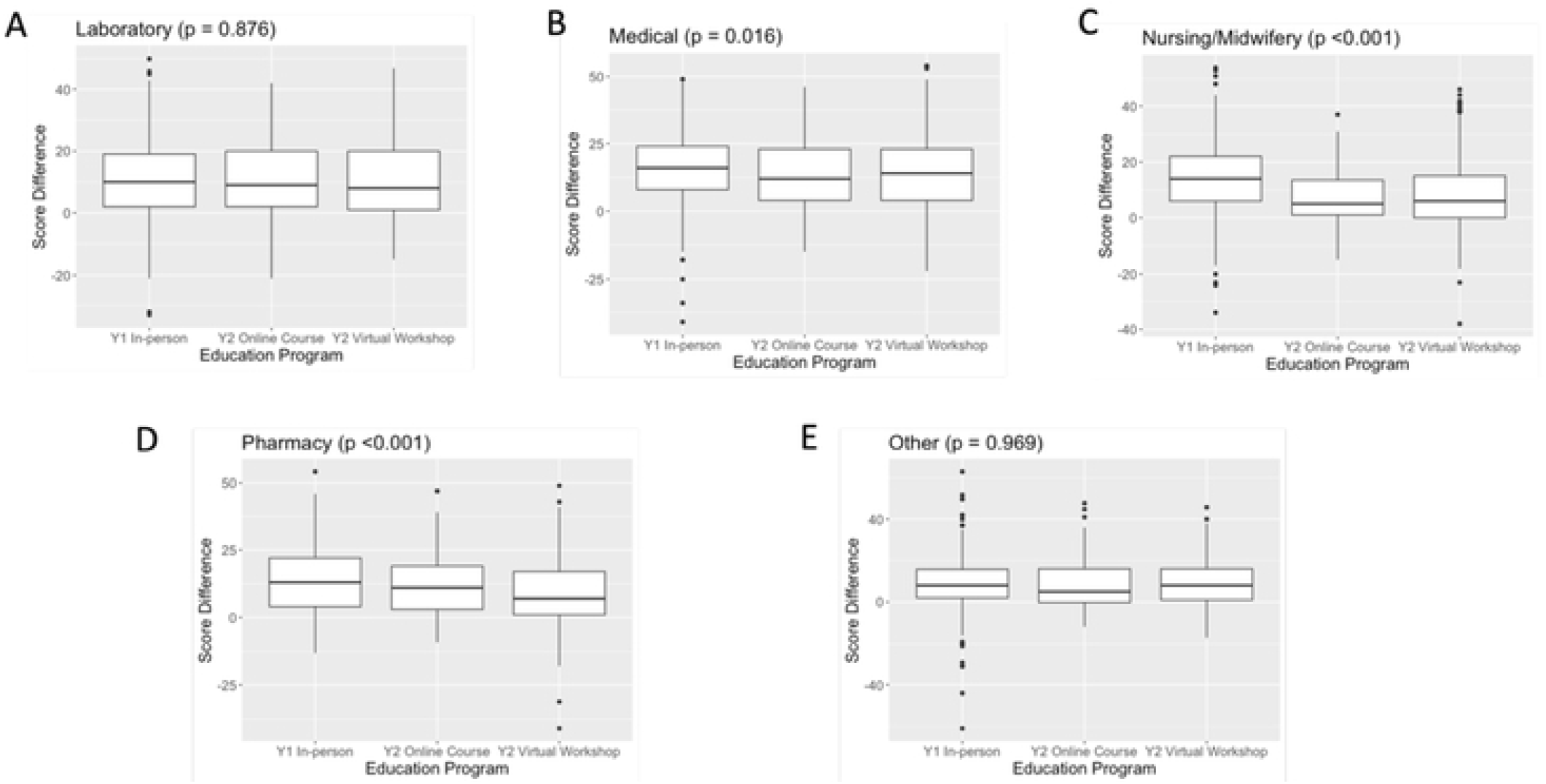
Total confidence score difference by education program for Laboratory (A, n = 365), Medical (B, n = 902), Nursing/Midwifery (C, n=1145), Pharmacy (D, n = 312), and Other (E, n = 299).

## DISCUSSION

In this multi-country capacity-building intervention, which included over 13,000 learners from diverse cadres and clinical settings, in-person and different online educational strategies were demonstrated to be similarly effective means of training learners in inter-professional HIV care across SSA. Moreover, the transition from in-person to online and remote training that occurred because of the COVID-19 pandemic, did not have a major deleterious impact on learner outcomes, as demonstrated by the gains in both knowledge and confidence across all three educational programs and for all types of learners. Nonetheless, gains in knowledge and clinical confidence were greater among in-service learners in Y1 compared to learners in Y2. These findings have several important programmatic and policy implications, as outlined below.

Firstly, it was notable that while the magnitude of gains in technical knowledge and clinical confidence were smaller among learners who participated in the synchronous Virtual Workshop and blended Online Course compared to those in the in-person training. Across all domains of confidence assessed, mean improvements in subjective confidence were greatest among learners who participated in the in-person program, regardless of cadre and career stage. These findings validate other research demonstrating the importance of online tools as satisfactory for acquiring knowledge, but inferior modalities for acquisition of clinical skills or technical confidence.(14, 15) While these findings underscore the importance of in-person learning for inter-professional training, they should be weighed against the relative merits of online remote learning modalities. For example, online learning tools are likely to be more affordable in many parts of SSA, especially given that learners can participate even from remote or inaccessible geographic locations.^10^ Recent studies in SSA have highlighted substantial opportunity costs associated with in-person training programs for in-service learners that are potentially obviated by distance learning modalities.(7, 8, 16) These benefits notwithstanding,(17) more research is warranted to ensure that online capacity building interventions are used effectively, and in combination with traditional in-person strategies, to foster clinical confidence and inter-professional collaboration. It is notable that confidence gains were greater for learners who participated in the synchronous Virtual Workshops compared to the blended Online Course format, highlighting that even when in-person workshops may not be feasible or affordable, strategies to facilitate real-time learning using online video-streaming platforms, can be leveraged, even in resource-variable settings in SSA.

Secondly, this analysis underscores how educational technologies can play a critical role in addressing inequities in access to training opportunities in SSA. Despite the profound, disruptive impacts of the COVID-19 pandemic, both the online synchronous and blended programs enabled more diverse and inclusive approaches to inter-professional HIV training for in-service learners. Learners in Y2 were more likely to be female and from more diverse cadres than learners in Y1. These findings are in contrast with other data highlighting how the COVID-19 pandemic revealed and increased pre-existing inequities in professional development opportunities for healthcare professionals in LMICs,(2, 18, 19) Although many health professions training institutions in SSA lack access and capacity to use digital technologies to deliver HIV training(20) our results affirm the critical role that online training interventions can play in advancing professional development opportunities especially for those healthcare professions historically underserved by capacity building interventions.(21)

Thirdly, this study highlights the importance of adaptive and resilient educational strategies in response to an emergent infectious disease threat. Although very few of the partner institutions involved in the program had adopted online learning tools before COVID-19, the pandemic accelerated a shift to digital technologies in health professions education even in countries with limited digital infrastructure. Moreover, use of digital platforms allowed learners in diverse settings to acquire new knowledge rapidly and effectively. While further research is critical to better understand how to optimally leverage digital tools in combination with more immersive in-person teaching formats, our study offers an African-specific blueprint for how to rapidly upskill the healthcare workforce, even in the setting of a major public health emergency.

The study has several limitations. Firstly, data is from an HIV-specific training program, and evaluative data limited to learners who completed the registration survey and assessment questions. Further research is warranted to determine if lessons learned from the analysis are applicable to other disease-specific capacity building interventions. Additionally, our assessment was limited to knowledge and confidence, as data regarding clinical outcomes was not available. Since different health profession training institutions participated in the program in Y1 and Y2 (in part due to digital literacy and internet infrastructure), we were unable to compare learner outcomes between Y1 and Y2 within individual institutions or country. Nonetheless, we assert that the aggregate findings have broad applicability to diverse Africa settings. Finally, institutions in Y2 were different from Y1 since participation was limited by digital literacy and Internet infrastructure. In Y2, we only included information from learners that were able to participate in the digital modalities highlighted here; the study does not address barriers faced by learners unable to access online training programs in SSA. These limitations highlight the ongoing need to support the development of local technological infrastructure for online education across SSA.

## Summary

In conclusion, this study highlights the relative impact of different educational strategies for enhancing HIV knowledge and confidence among clinicians and pre-service learners in SSA. While in-person learning resulted in greater increases in knowledge and confidence, remote training modalities were demonstrated to be effective. These results are important for decision-makers as they choose strategies to improve the HIV programs in SSA, especially since they highlight how digital learning solutions ensured increased access to professional development opportunities for frontline clinical providers in diverse African settings. Further research is warranted to evaluate the impact of digital learning tools in SSA, including assessing feasibility and cost utilizing implementation science frameworks.

## Data Availability

Data are available in a public, open access repository. Extra data can be accessed via the Dryad data repository at http://datadryad.org/ with the doi:10.7272/Q6WQ021N.

https://doi.org/10.7272/Q6WQ021N

## Notes

**Role of Funding** This work was supported with funding from Health Resources and Services Administration

**Conflict of Interest Statement** We declare that we have no conflicts of interest.

### Competing Interest Statement

The authors have declared no competing interest.

### Author Declarations

The design of the training program, including the topics covered and the format of the training, was informed by input from focus-group discussions with patient groups, learners (both pre-service and early career professionals) and HIV educators from a variety of settings in SSA, and has been previously described. Assessment tools, evaluating learners knowledge and confidence, were also piloted with a subset of multidisciplinary learners before the full program was launched. All learners were given access to their pre and post score test results, via the program’s website. In addition, aggregate, site-level evaluation data were also posted on the program’s website. The protocol for this project was reviewed and approved by the University of California, San Francisco’s Institutional Review Board (IRB) in San Francisco, California. Verbal consent was required at the time of participation in the study as approved by the IRB (protocol #: 19–28,447).

